# Legionnaires’ disease on the rise in Switzerland: A denominator-based analysis of national diagnostic data, 2007-2016

**DOI:** 10.1101/2020.05.27.20114355

**Authors:** Fabienne B. Fischer, Claudia Schmutz, Valeria Gaia, Daniel Mäusezahl

## Abstract

**Purpose:** The risk of Legionnaires’ disease (LD) is suggested to increase, but the global burden of disease is unknown due to lack of appropriate diagnosis and surveillance systems. In Switzerland, the number of LD cases, captured by the National Notification System for Infectious Diseases, has more than doubled since 2008. The aim of this study is to evaluate disease surveillance data using denominator data i.e. the number of tests performed for *Legionella*. spp.

**Methods:** We collected the testing data for *Legionella* spp. of 14 Swiss diagnostic laboratories and calculated the positivity rate, which is the rate of the number of positive tests to the number of tests performed.

**Results:** The number of positive tests increased proportionally to the number of tests performed; hence the positivity remained stable. However, the cause of the increase in test volume is unclear and has a large impact on the interpretation of the positivity curve. Further, the test outcome was found to be dependent on regional determinants and the diagnostic method applied.

**Conclusion:** In concert with the lack of understanding if and at which stage LD is considered in current case management of pneumonia patients, it is difficult to interpret heterogeneities in incidence or underestimation in Switzerland.

## INTRODUCTION

*Legionella* spp. is the cause of a group of diseases termed “legionellosis” ranging from mild and self- limiting Pontiac fever to potentially fatal Legionnaires’ disease (LD), characterised by pneumonia [1, 2]. Infection with *Legionella* spp. occurs through inhalation or aspiration of contaminated water or aerosols. In recent years, person-to-person transmission was also suspected [3]. Cases can occur sporadically, in clusters and large outbreaks.

Although *Legionella* spp. occurs worldwide, the global burden of disease is unknown due to the lack of appropriate diagnosis and/or surveillance systems in many countries. In Europe in 2017, 1.8 cases per 100’000 population were estimated, corresponding to 9’238 cases in total. In the same year, the US reported 7’500 cases, corresponding to 2.3 cases per 100’000 population [4, 5]. Case numbers have been increasing in European countries and the US in the past years.

In Switzerland, infections with *Legionella* spp. need to be reported to the National Notification System for Infectious Diseases (NNSID), which is managed by the Federal Office of Public Health (FOPH), since 1988. While all laboratory-confirmed infections are notifiable, only LD cases – cases with pneumonia – are considered as confirmed or probable cases, which are reflected in numbers published in official statistics. The case numbers continuously increased from 219 in 2008 to 464 cases in 2017 [6].

The increase of LD cases in Switzerland, the rest of Europe and the US, is not well-understood. It has been hypothesised that the increase in incidence is due to augmented susceptibility in the population, climate change or changes in energy policies [4, 6]. Common risk factors for LD are age >40 years, being male, tobacco smoking, traveling abroad or having chronic conditions, e.g. diabetes mellitus or a compromised immune system [7, 8]. Further, several studies link weather and climate, namely warm and humid conditions, to LD incidence [9-12]. Efforts in energy saving resulting in recommendations to lower temperature thresholds of potable warm water, could have the drawback to promote conditions which favour *Legionella* spp. proliferation [13].

Conversely, the increase in case numbers could also be an artefact. Increased awareness of physicians could lead to increased testing and hence, to more cases found. The incidence of legionellosis is generally thought to be underestimated; a study from Germany in 2008 estimated about 15’000 to 30’000 cases of sporadic LD annually [14]. Improvements in diagnosis and surveillance could lead to higher but more accurate case numbers [15, 16].

To evaluate the effect of changes in test numbers and diagnostic procedures on the notification numbers in Switzerland, we collected testing data of 14 Swiss diagnostic laboratories between 2007 and 2016. Using this data, we calculated the positivity of *Legionella* spp. testing, emphasising on temporal trends, and assessed the determinants for a positive test outcome.

## METHODS

The methods of a positivity study have been described in detail elsewhere^1^. In brief, we collected testing data from 14 Swiss diagnostic laboratories. The laboratories were selected in 2016, based on providing most LD notifications in the 10 years prior.

We collected data on all tests performed for *Legionella* spp. regardless of test outcome, between January 2007 and December 2016. Information requested included date of test, test result (binary 0=negative; 1=positive), diagnostic test method, sample material used, patient identification number, and patients’ date of birth, sex and canton of residence. We excluded tests of patients with residency outside of Switzerland, inconclusive test results, duplicated entries as well as “repeated tests”. Repeated tests were defined as more than one test performed per patient and disease episode. The definition of a disease episode was complex given the laboratory data available; the process is described in the supplementary material (see Online Resource 1 for details).

The analysis was planned *a priori* and was conducted using STATA 15 (StataCorp., USA).

We use the term positivity as the proportion of the number of positive tests to the total number of tests performed for *Legionella* spp. [17, 18]. The positivity was calculated for different age and sex groups, test methods, sample materials, spatial (region and laboratory) and temporal (annual and seasonal) trends. The main outcome, the annual positivity, was age- and sex-adjusted using direct standardisation with the sample population (2007-2016) as reference population.

We used mixed effect logistic regression to account for clustered data to analyse the determinants for a positive test result. The significance level was defined as α= 5%. Univariable logistic regression was used to test the association between test result and test year, season, time trend, sex, age group, laboratory, test method, sample material and greater region. Season was modelled using a sine and cosine function with an annual period. The time trend was a continuous variable combining test month and test year. The age groups were based on categories (standard in ECDC publications) but we used a higher level of differentiation in older people, due to the known risk factors for LD. The greater regions correspond to the Nomenclature of Territorial Units for Statistics (NUTS)-2-level. Categories with most observations were chosen as reference categories, except for the seasonality (first month of the year).

We constructed two multivariable mixed effect logistic regression models, including both the variables sex, age group, season, time trend, and test method. One model included the region and the other the laboratory as random effect. This partition was necessary due to collinearity and bias between the two variables and the outcome variable.

### Ethical statement

The study was conducted under the Epidemics Act (SR 818.101). The data, provided by laboratories, were anonymised for analysis. Other data (notification data, population statistics) are publicly available from the FOPH or the Swiss Federal Statistical Office.

## RESULTS

### Data received

The 14 laboratories provided a total of 154’851 observations, including 2’808 positive tests. Three laboratories could not provide data for the entire study period (2007-2016) due to changes in their laboratory information system and data storage.

### Exclusion

Applying pre-defined exclusion criteria (residence outside of Switzerland, inconclusive test results, tests performed outside of the study period), we excluded 6’721 observations (134 positives and 968 with an inconclusive or missing test result). Additionally, 762 (13 positive) entries were excluded, with information on either sex or age was missing.

We excluded 7’287 duplicates (412 positives) from the dataset. It was further decided to exclude all serological tests. In total 2’558 (1.8%) serological tests (108 positives) were performed (see Online Resource 2 for details). Lastly, 13’196 repeated tests (383 positives) were excluded. The final dataset comprised 126’422 (1’638 positive) observations.

### National Notification System for Infectious Diseases

We compared the number of positive test results in our dataset to the NNSID notification numbers as notified by our selected laboratories. As noted above, the published notification numbers only reflect LD cases while the positive test results in our dataset reflect all legionellosis cases. The biggest difference in numbers was observed in 2009 with a relative difference of 27.6% (91 LD cases in the NNSID compared to 141 positive test results in our dataset, Fig 1). The average relative difference was 11.1%. Generally, the annual case number from all participating laboratories combined was higher in our dataset than in the NNSID data.

**Fig 1.**
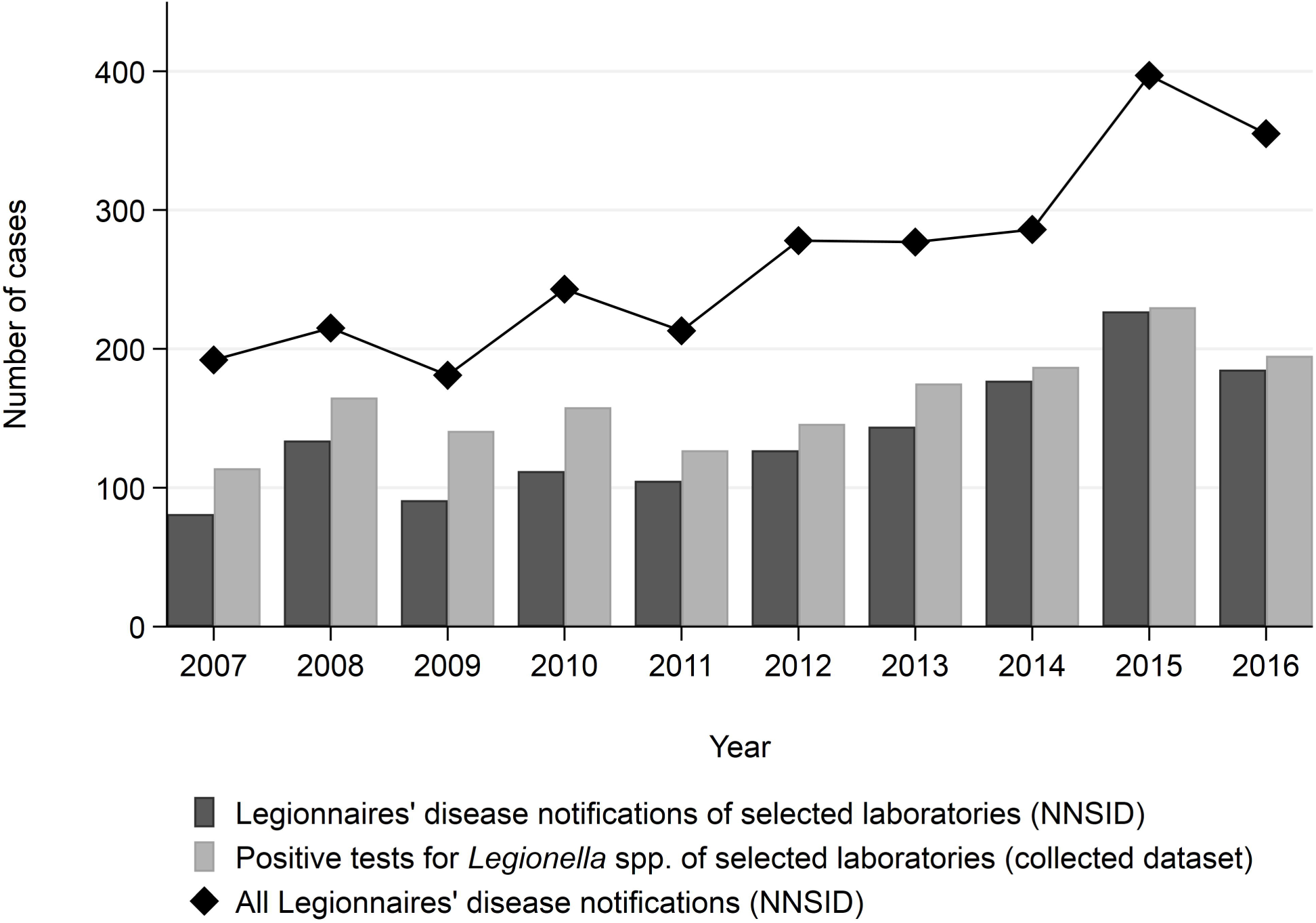
Legionnaires’ disease notifications. Number of Legionnaires’ disease (LD) notifications of the 14 selected laboratories as reported in the Swiss National Notification System for Infectious Diseases (NNSID) and the number of positive tests of the selected laboratories, as well as the total number of LD notifications reported in the NNSID per year, 2007-2016, Switzerland

The LD cases notified to the NNSID from the 14 selected laboratories account for 54% of all notified cases nationwide between 2007 and 2016. This proportion remained constant across the years.

### Positivity

The number of tests performed increased by 131% from 7’366 in 2007 to 17’027 in 2016 and the number of positives by 71% from 114 to 195 (Fig 2a). The yearly age- and sex-adjusted positivity decreased marginally from 1.5% to 1.1% (Fig 2b).

**Fig 2.**
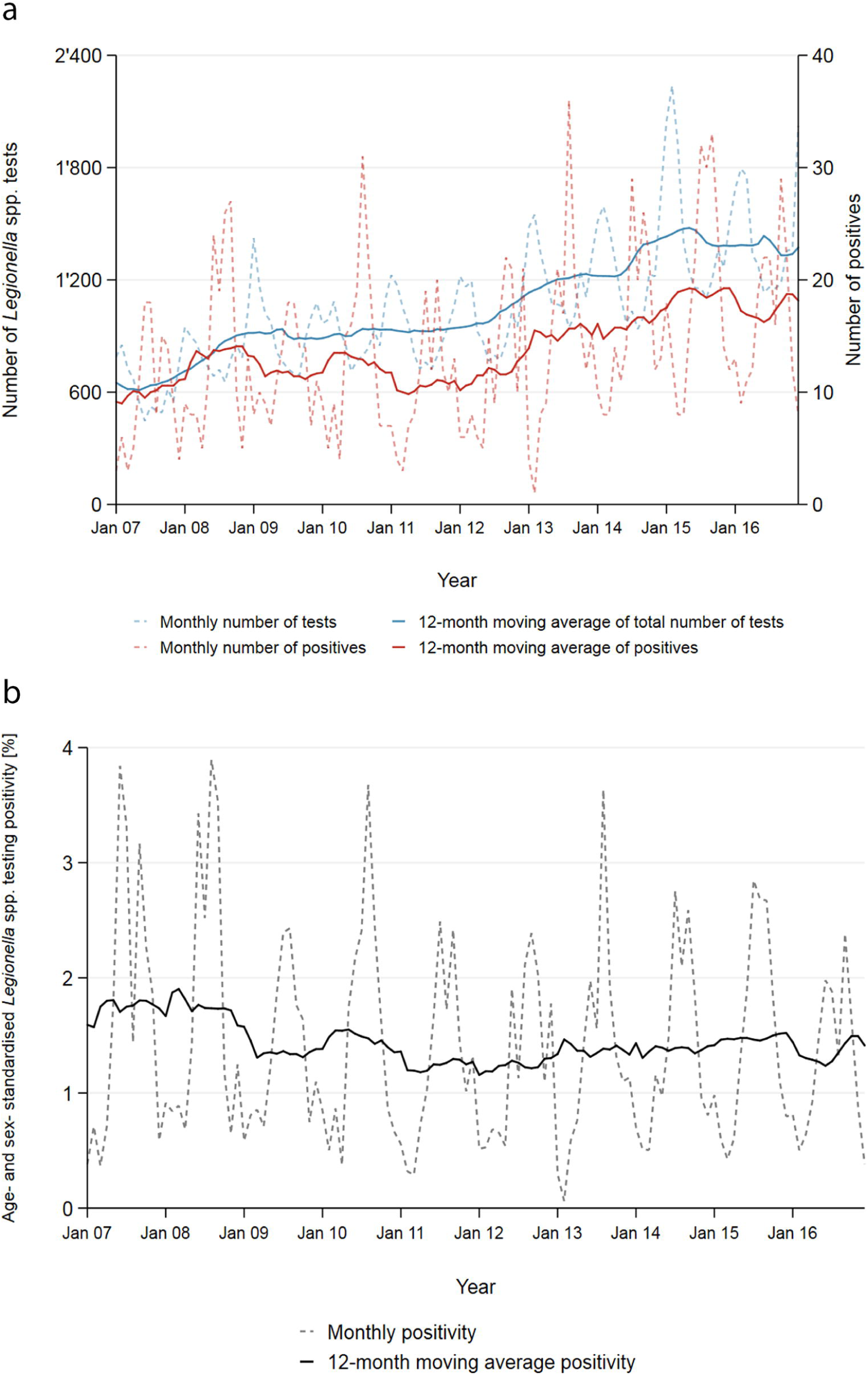
Time trend in test volume, cases and positivity. a: Twelve-month moving average (solid lines) and monthly (dashed lines) number of *Legionella* spp. tests performed and number of positive tests, for the entire study period (2007-2016) by 14 diagnostic laboratories in Switzerland. b: Twelve-month moving average (solid line) and monthly (dashed line) age- and sex-standardised positivity of *Legionella* spp. testing, Switzerland, 2007-2016

Across all years, the positivity started increasing in May and peaked in August and September reaching 2.6%, then decreased in October to reach an all year low in February with 0.5%. The seasonality of the positivity is a direct result of the contrasting seasonality of the number of tests performed and the number of positive test results obtained (Supplementary Fig 1). Most tests were performed during the winter months; on average 62% more tests were conducted in February than in August. Conversely, more than three times as many cases were reported in September compared to February.

The seasonality persisted across all age groups, both genders and all regions. It is most strongly reflected in tests performed by UAT. PCR and culture-based tests do not show any clear seasonal pattern for the number of tests performed and the number of positive cases, also explained by small numbers.

### Gender and age

The positivity varies strongly by gender and age group. Males have an overall higher positivity compared to females (1.6% to 0.9%). The positivity increases with age and is highest among 45-64 year olds (2.5% for males and 1.3% for females) and then decreases gradually again; this pattern is similar for both genders (Supplementary Fig 2a). The positivity of males aged 5-14 years old is the only exception to the bell-shaped distribution with a positivity of 1.4%. No female in the age groups “0-4” and “5-14” was tested positive.

The majority of patients tested were males (57.9%, N=73’224). This proportion remained stable across the study period (2007-2016). The overrepresentation of males in the tested population was seen in all age categories, except in the oldest (85+ years old), where 48.7% of all patients were male (chi-square test: p<0.01, Supplementary Fig 2b).Overall, most tests were performed in the age group of 75-84 year olds (25.6%, N=32’349), closely followed by the age group of 45-64 year olds (24.5%, N=30’956). Least tests were performed in the age groups of infants (0-4), adolescents (5-14) and young adults (15-24) with 0.3%, 0.4% and 2.3% respectively. During the study period, the age distribution of tested patients remained similar and the median age increased only marginally from 69 years old in 2007 to 71 in 2016 (Kruskal-Wallis test: p<0.01).

The difference in sex distribution was small but statistically significant for all greater regions (range 57.5% to 60.4% males, chi-square test: p<0.01) and slightly more variable between laboratories (53.3% to 64.4% males, chi-square test: p<0.01). Similarly, the median age only differed marginally, but significantly between regions (range of medians 68-73 years old, Kruskal-Wallis test: p<0.01) and more strongly between laboratories (range 59-74 years old, Kruskal-Wallis test: p<0.01).

### Regional differences

Of the 14 laboratories in our dataset, 11 were hospital laboratories accounting for 86.2% (N=109’016) of all observations included in this analysis. However, the three private laboratories may also perform diagnostics for hospitalised patients. The laboratories performed diagnostics mainly for patients with residency in proximity to the laboratory site. Therefore, the variable “laboratories” is correlated with the variable “greater region” (correlation coefficient r=0.12, p<0.01, Supplementary Fig 3). Hence, any information on regions is heavily influenced by the selection of laboratories.

The positivity in the greater regions across all years ranged from 0.9% in “Northwestern Switzerland” to 2.4% in the region “Zurich” (Fig 3). The positivity for all regions decreased from 2007 to 2016 except in “Northwestern Switzerland”, where there was a relative increase of 44%. The positivity fluctuates throughout the years, most notably in “Zurich” (range 1.1% to 4.5%).

**Fig 3.**
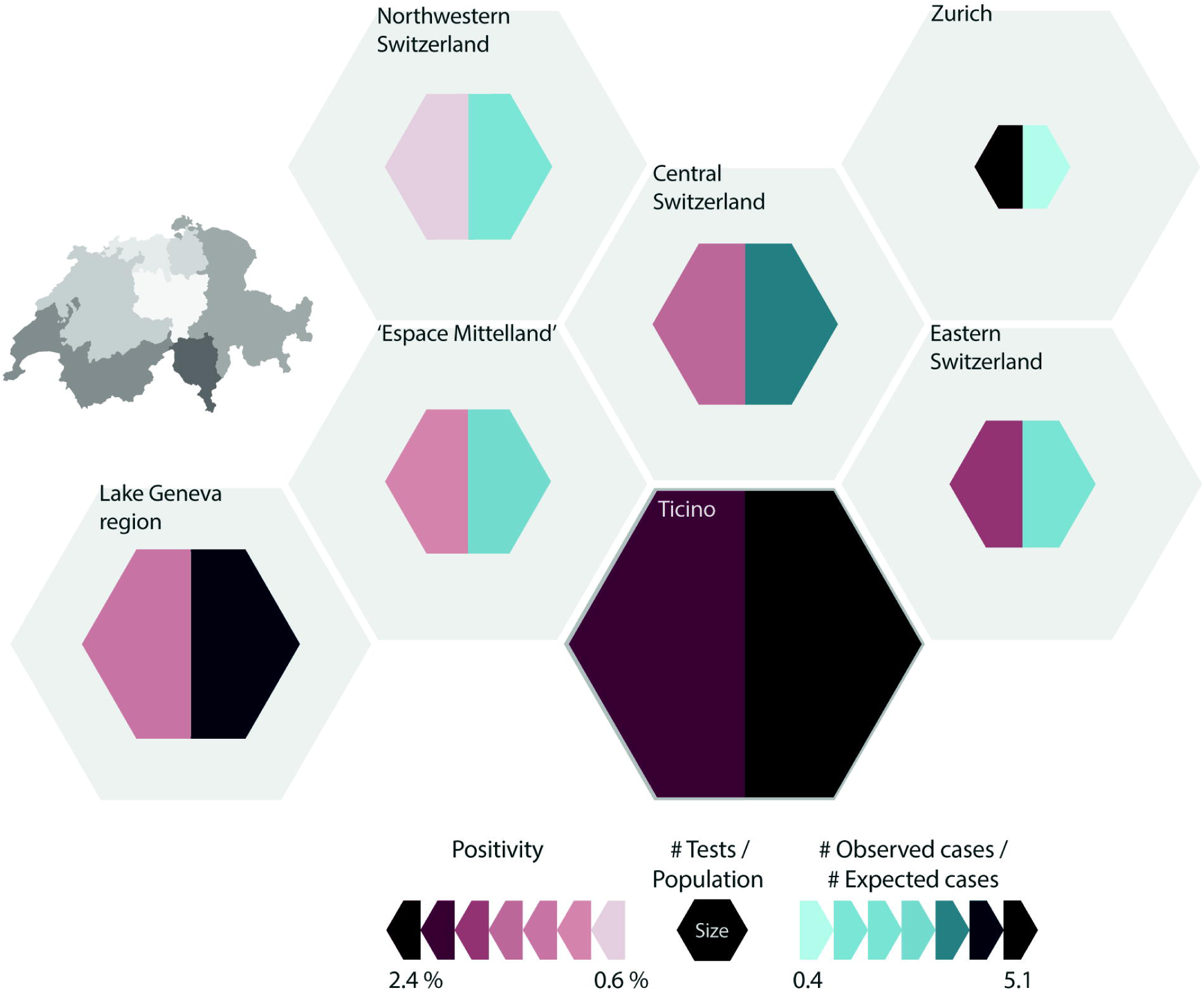
Distribution of test volume, cases and positivity across Switzerland. Representation of the seven greater regions of Switzerland (displayed as grey area) of the positivity for *Legionella* spp. testing (colour of left part of the smaller hexagon), the number of tests performed in relation to the resident population (size of smaller hexagon) and the ratio of the number of observed cases to expected cases (colour of right part of the smaller hexagon), based on the testing data of 14 Swiss diagnostic laboratories (2007-2016)

Over the entire study period (2007-2016), most tests were performed in the “Lake Geneva” region, followed by “Espace Mittelland” and “Ticino”. The least amount of tests was reported from the region “Zurich”. In relation to the average population of the regions (2007-2016), much more tests were conducted in “Ticino” with 6’493 tests per 100’000 population compared to “Zurich” with 377 tests per 100’000 population (Fig 3). The average for all greater regions was 2’028 tests per 100’000 population.

The number of tests performed increased in all regions between 2007 and 2016. The biggest relative increase (129 in 2007 to 1’698 in 2016) was observed for “Northwestern Switzerland”, followed by “Espace Mittelland” (544 to 3’055) and “Eastern Switzerland” (455 to 1’332). “Zurich” had the smallest relative increase (536 to 585). This distribution remained stable, even when disregarding the laboratories not providing data for the entire study period.

### Diagnostic methods and sample material

The process of exclusion of repeated tests could already provide first insights on the diagnostic procedures used in the laboratories. In the raw dataset 7.7% (N=10’809) of all patients were tested at 6least twice during the same disease episode; 4.3% (N=6’022) of the patients were tested more than once on the same day. Omitting tests performed on the same day - as order of test could not be assessed - 3.4% of all UATs, 14.8% of all culture-based tests and 14.7% of all PCR tests, were excluded as repeated tests. The positivity among the repeated tests was 2.9%.

All results henceforth stem from the analysis of the cleaned dataset (omitting repeated tests). The positivity of *Legionella* spp. tests performed using UATs was 1.3%, using culture-based tests it was lower (0.8%) and using PCR higher (3.1%). The positivity of UATs varied based on the exact test used (Fisher’s exact test: p<0.01): The UATs from lowest to highest positivity were, Binax™ *Legionella* Urinary Antigen EIA (Alere) (0.9%), BinaxNOW® *Legionella* ICT (Alere) (1.2%), Biotest *Legionella* Urinary Antigen Enzyme Immunoassay (EIA, Biotest) (1.7%) and Sofia *Legionella* Fluorescent Immunoassay (FIA, QUIDEL) (2.2%).

The positivity of UATs decreased during the study period from 1.6% in 2007 to 1.1% in 2016. The positivity of culture-based tests remained below 1% except for three years (2008: 1.5%; 2012: 1.6%; and 2014: 1.4%). The positivity of diagnostic tests using PCR increased gradually since 2011 from 2.8% to 4.8%.

The majority of diagnostic tests performed were UATs with 90.1% (N=113’863) followed by culture-based methods (6.6%, N=8’373) and PCR (3.3%, N=4’169). This distribution remained stable at large between 2007 and 2016. Only PCR slightly gained importance (0.8% in 2007 to 2.5% in 2016) at the costs of UATs (92.4% to 88.3%).

UATs performed were mostly BinaxNOW (71.4%), Binax (11%), Biotest (10.1%) and Sofia *Legionella* FIA (7.6%). The Sofia *Legionella* FIA test was introduced only in 2014 and increased its market share since to 28.6% of all UATs in 2016. For BinaxNOW market shares decreased from 75.7% of all UATs in 2007 to 52.3% in 2016.

Almost all of the nine laboratories performing PCR used a different type of test. Four laboratories reported to outsource PCR diagnostics to other laboratories and could, therefore, not provide detailed information. Three laboratories had communicated to use respiratory multiplex PCR panels, however, for two of three the distinction between single and multiplex PCR could not be made in our dataset (personal communication, May-July 2017). Due to this heterogeneity and lack of accuracy, we did not further quantify the different types of PCR tests performed.

As the test method is dependent on the laboratories and their diagnostic procedures, the variable “method” is correlated with the variable “laboratory” and therefore also with “region”.

Twelve of the 14 laboratories predominantly or exclusively performed UATs. One laboratory performed 76.3% PCRs and another 79.3% culture-based diagnostics. The proportion of PCR increased in the former between 2007 and 2016 replacing UATs while in the latter, the proportion of culture-based tests and UATs increased replacing PCR.

UATs comprise at least 80.6% of all tests performed in all greater regions. The biggest proportion of culture-based tests was performed in “Espace Mittelland” (11.5%), “Lake Geneva region” (9.8%) and “Northwestern Switzerland” (8.9%). Most PCR tests were performed in “Northwestern Switzerland” (10.5%) and “Lake Geneva region” (3.2%). In four of the seven regions, the diagnostic methods used over the years did not change.

### Determinants for a positive test result of *Legionella* spp

The univariable model showed a significantly increased odds ratio (OR) for a positive test outcome for the test years 2007 and 2008 compared to the latest test year 2016. The time trend variable showed a marginal downward trend, with a rounded OR of 1 (exact OR 0.998, CI 0.9970-0.9998, p=0.03). Further, all calendar months from May to December had significantly increased odds for a positive test outcome compared to February. The highest odds were calculated for August and September (OR 4.02, p<0.01 for both).

Females were almost half as likely as males to be tested positive for a *Legionella* spp. infection (OR 0.56, p<0.01). Compared to the reference group of 75-84 year olds, the age groups “15-24” and “85+” had significantly decreased odds for a positive test outcome (OR 0.41, p<0.01 and OR 0.76, p<0.01) and the age groups “45-64” and “65-74” showed increased odds (OR 2.1, p<0.01 and OR 1.36, p<0.01).

“Northwestern Switzerland” showed 20% lower probability for a positive test result compared to the “Lake Geneva” region (OR 0.81, p=0.04), while “Zurich” had more than double the odds and “Ticino” a 50% increased chance for a positive test result (OR 2.2, p<0.01 and OR 1.47, p<0.01).

Culture-based tests had lower odds for a positive test outcome compared to UATs (OR 0.63, p<0.01). In contrast, PCR tests had 2.5-fold increased odds for a positive test (OR 2.47, p<0.01).

The univariable regression using the sample material as explanatory variable was stratified by culture-based tests and PCR. For culture-based tests using material obtained through paracentesis or using sputum showed the highest OR (OR 10.32, p=0.03 and OR 5.12, p<0.01). Using PCR, material obtained through paracentesis (OR 4.47, p=0.05) or swabs (OR 3.91, p<0.01) or using sputum (OR 3.24, p<0.01) had elevated odds for a positive test outcome.

Fig 4 shows the ORs for different UATs before and after inclusion of “laboratory” as a random effect. Univariable models including other variables showed no significant effect on the ORs and are therefore not shown.

**Fig 4.**
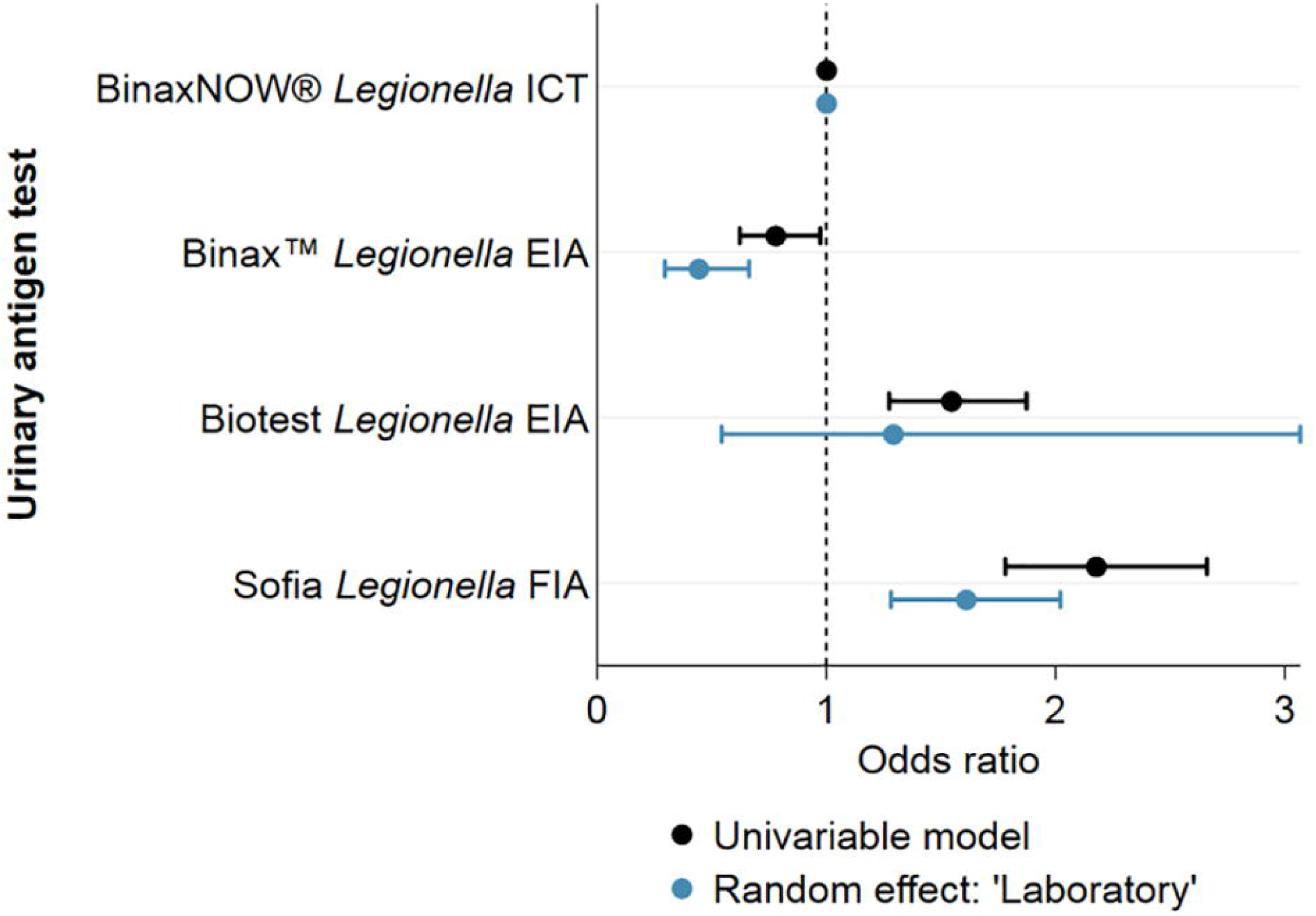
Differences in positivity across UAT test kits. Univariable regression results with and without random effect on “laboratory” for the outcome of having a positive test result for *Legionella* spp. in Switzerland, 2007-2016

Both multivariable mixed-effect logistic regression models (with the inclusion of “region” or “laboratory” as random effect, respectively) are shown in Fig 5 together with the results of the univariable models. The estimates are comparable for all variables. The marginal but statistically significant negative OR for the time trend-variable, however lost its statistical significance in both multivariable models.

**Fig 5.**
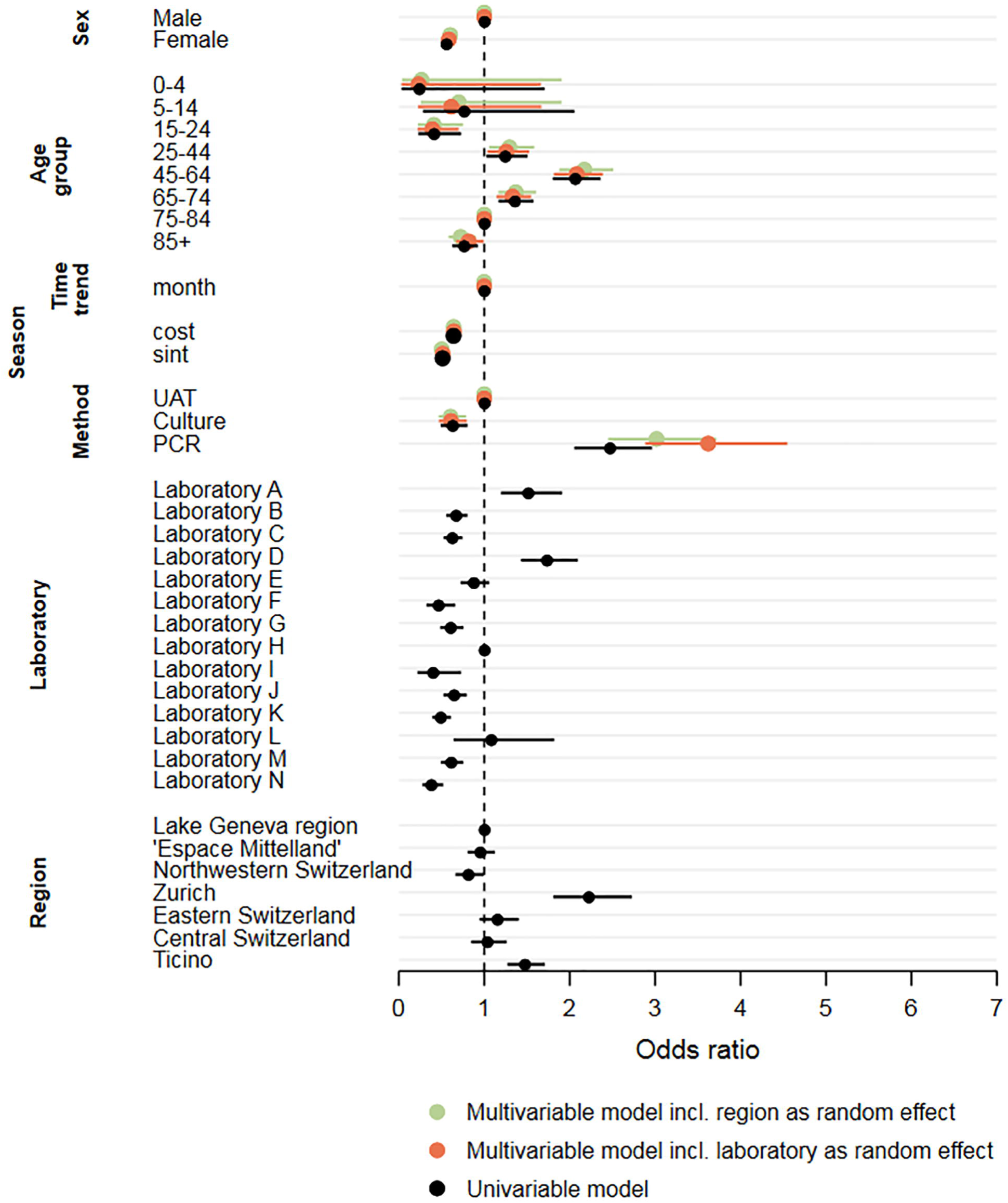
Determinants for a positive test result for *Legionella* spp. Multivariable mixed-effect logistic regression compared with the univariable regression results for the outcome of having a positive test result for *Legionella* spp. in Switzerland, 2007-2016

## DISCUSSION

We collected the testing data of 14 Swiss diagnostic laboratories and calculated the positivity rate, i.e. the relationship of the number of positive tests to the number of tests performed to investigate the increase observed in case numbers of LD in official disease surveillance.

### Time trend in positivity 2007-2016

The number of *Legionella* spp. tests performed increased more strongly than the number of cases found, resulting in a marginally decreasing positivity between 2007 and 2016 from 1.5% to 1.1%. However, no temporal trend was found in the multivariable regression models. The strong increase in test numbers for *Legionella* spp. cannot be explained given that contextual information on health-seeking, test behaviour of physician and on diagnostic methods and procedures applied by laboratories are essential for a correct interpretation of trends in positivity.

We hypothesise that changes in the diagnostic methods influenced the number of tests performed: Especially the introduction of the urinary antigen test (UAT) revolutionised the diagnosis of legionellosis. In 2015, 78.2% of all LD cases in Europe were detected using UATs [19]. In Switzerland, UATs have been introduced to the routine diagnostic in 1997 and are now predominantly used [20]. However, the UAT is unlikely to have influenced the most recent increase in test numbers, as the introduction of this test occurred almost 20 years ago and the proportion of UATs performed remained stable or declined during the study period. Therefore, changes in testing behaviour of physicians, health-seeking behaviour of patients, prevalence in risk factors, and in disease frequency need to be considered to explain the increase in test volume.

Symptom-based testing explains the inverse seasonality in the number of tests performed and the number of cases found. Community-acquired pneumonia (CAP) peaks during the winter months but is predominantly caused by agents other than *Legionella* spp. [21]. Therefore, the testing volume is higher in winter than in summer even if the physicians are aware of a summer peak for *Legionella* spp. [22].

The Swiss Society of Infectious Diseases (SSI) provides guidelines for the management of CAP which were adapted from European guidelines. The guidelines state that microbial testing is not indicated in primary care settings and even in hospital settings *Legionella* spp. testing may only be useful for selected risk patients based on clinical or epidemiological features [23]. These recommendations leave room for interpretation and can, thus, be applied differently by the treating physician, depending on his/her awareness of LD and knowledge of its epidemiological and clinical features. Failure to diagnose LD has previously been attributed to a lack of awareness relating to LD [24]. Heightened awareness of physicians and consideration of LD in their differential diagnosis of patients presenting with pneumonia would lead to more LD tests ordered over time. However, there is a lack of information on adherence to the CAP guidelines, the awareness level among Swiss physicians and the case management of LD in Switzerland.

An increasing number of patients in Switzerland seek care for non-urgent or non-life-threatening conditions at emergency departments rather than at primary care level [25]. These “new” patients presenting at the emergency department could contribute to a higher number of LD tests conducted: according to SSI guidelines, microbiological investigation of pneumonia is recommended earlier in the hospital compared to the primary care setting. Increased awareness and change in health-seeking behaviour as potential causes for increased testing are independent of disease incidence, but rather represent a shift in test practices. More cases are found if a larger part of the population is being screened for LD, hence, reducing the extent of underestimation.

However, an important alternative explanation for the increase in test volume is that actually more ill patients present with signs and symptoms of LD (i.e. pneumonia). In this scenario, the increase in test numbers would be explained by an increase in incidence rather than a decrease in the extent of underestimation. According to the “medical statistic of hospitals” (“Medizinische Statistik der Krankenhäuser”) published annually by the FSO, the number of hospitalised patients with pneumonia as recorded “main diagnosis” in over 14-year-olds has increased by one third between 2007 and 2016, while the number decreased for patient younger than 15 [26]. Hence, these statistics support the hypothesis of increased pneumonia incidence leading to higher test volumes.

The investigation of positivity curves seems to bring most insights in disease and surveillance trends if changes in diagnostic methods at laboratory level occurred and are assumed to cause changes in notification numbers. For example, the introduction of multiplex PCR was suspected to impact on EHEC surveillance (Reference EHEC positivity paper, Fischer *et al*. 2018 [under review]). However, we found that *Legionella* spp. testing was not subject to such a significant change. Additionally the lack of information and understanding of the trajectory from health-seeking to LD diagnosis does not allow conclusive interpretation of the 10-year-trend in positivity rate.

The number of reported LD cases is not only rising in Switzerland but also in the EU/EEA countries, which are members of the European Legionnaires’ Disease Surveillance Network (ELDSNet) and in the US [4, 26]. Yet, Switzerland had the highest notification rate per 100’000 population in 2017 (5.8), followed by Slovenia (5.7), Denmark (4.8) and Italy (3.3) and the second largest increase in notification rate between 2013 and 2017 [4, 27]. However, we are not aware that data from these national surveillance systems have been evaluated considering denominator data. Furthermore, notification rates are heavily influence by the health system itself and hence, comparability between countries is limited.

### Male and elderly people at risk

We found that men were more often tested for *Legionella* spp. than women and positivity was significantly higher for males than for females. This suggests that the higher case numbers for men are actually due to a higher incidence in the male population or a diverging health-seeking behaviour rather than a more thorough testing due to male sex being a known risk factor [28].

Regardless of gender, adults over 25 years showed an increased positivity, peaking at 45-64 year of age and declining again in older age groups. The difference in positivity for the middle aged patients (25-64) compared to the older patients (over 65) is likely due to a more thorough testing approach for the older patients compared to younger patients (testing elderly earlier, presenting with less severe acute respiratory infections). The difference in test volume could also be due to the knowledge that older age is a risk factor for LD or CAP being more prevalent in older age [6, 8, 29].

### Regional differences across Switzerland

It is difficult to estimate regional differences based on our data, as it is heavily dependent on the laboratories included in the study. The interaction of these variables (“laboratory” and “greater region”) is not straight-forward to assess. Apart from the collinearity of these variables, we found that the positivity of tests performed in one laboratory differed substantially depending on the residency of the patient, especially if the patient lived outside of the usual catchment area of said laboratory. We assume this is due to different pre-test probabilities for a positive test outcome, e.g. only the samples of immunosuppressed patients (with an assumed higher probability of an actual infection with *Legionella* spp.) would be sent to another laboratory for confirmation. However, it is impossible to control our dataset for these ‘outsourced’ testing. For this reason, we decided to construct two multivariable logistic regression models. These limitations should be kept in mind when interpreting the results on the greater regions.

The calculated positivity and the logistic regression models show heterogeneity across regions. The regions of “Zurich” and “Ticino” seem to identify more positive cases per number of tests performed. Additionally, the two regions show opposite test frequency: relative to the resident population, in “Ticino” a lot of people are tested for LD while the inverse applies for “Zurich” It is known that in “Ticino” essentially all inpatients with pneumonia are tested for *Legionella* spp. infection (personal communication, February 2019). Further, “Ticino” has the highest notification rate amongst all Swiss cantons, which might not only result from the highest testing volume, but from an actually increased incidence, as indicated by the increased positivity and suggested by other studies focusing on the impact of climate on regional level (2018)[12]. In contrast in “Zurich”, where the number of reported cases and case rate is similar to the national average (2008-2017), either testing is more targeted to the “correct” patients, resulting in a higher positivity rate or incidence is actually higher, but underestimated due to the small test number [6]. In “Northwestern Switzerland” the reporting rate is also on the national average, but the region has significantly decreased odds for a positive test outcome. The number of tests performed increased most strongly in this region. At the same time, it was the only region with an increasing positivity.

As has been mentioned before, the SSI guidelines are subject to interpretation of the health personnel and the level of adherence is unknown. Hence, who is tested is likely very heterogeneous across Switzerland. We know of two hospitals, that do systematic screening of all pneumonia patients admitted to the emergency ward (personal communication, June 2016).

### Heterogeneity in the diagnostic methods

Not all diagnostic test methods for Legionella detect the same pathogens and strains. The application of UAT is limited to *Legionella pneumophila* serogroup 1, culture-based techniques can detect all Legionella species, while PCR techniques can either detect only *L. pneumophila* or also all species, depending on the type of test. The positivity also varies depending on the test method used, on the application of the chosen test method, but also on the specific kind of test kit or manufacturer. Using PCR increased the odds of obtaining a positive test result significantly compared to UAT and cultures. This could be attributed to the higher sensitivity of PCR compared to UAT, but could also result from false-positives due to cross-reaction or contamination [30]. A systematic review comparing UAT and PCR found PCR to be preferable. However, PCR is limited by the availability of appropriate sample material from the patient [31]. Culture is still regarded as the gold standard, as it allows cultivation of strains but exhibits an overall lower sensitivity, which corroborates with our results [30]. However, in our study the differences in positivity between UAT, PCR and culture could also be attributed to different testing behaviours, rather than to features inherent to the test. In some hospitals/laboratories, PCR and culture-based tests may only be performed in high-risk patients (e.g. immunocompromised patients), which might affect the positivity and introduce bias.

Double-testing with different diagnostic methods is often seen as advantageous [31-33]. The European Study Group of *Legionella* Infections (ESGLI) further recommends that all samples positive by UAT should be retested after heat treatment of the urine for confirmation, unless the initial sample was already boiled [34, 35]. However, in our raw dataset, only 1 in 12 patients got tested at least twice during the same disease episode. UATs are most often used as a stand-alone test and only in 194 cases a positive UAT was repeated during the same disease episode. However, we need to consider the possibility that not all secondary tests for confirmation might be registered in the individual laboratory information systems and, hence, are possibly not reported.

Lastly, the choice of test kit manufacturer for the widely used UAT also influences the positivity of *Legionella* spp. testing: the Sofia *Legionella* FIA test has a significantly increased positivity compared to the most commonly used UAT BinaxNOW. In comparison, the former has a higher sensitivity, but also a lower specificity especially without heat treatment of the urine, which could lead to false positive results [36]. The Swiss national reference centre for *Legionella* (NRCL) recommends the heat treatment of all urine samples, if Sofia *Legionella* FIA is used. If the initial urine sample was not boiled, a confirmation test needs to be performed on positive samples. However, it is unclear if and how many laboratories adhere to these recommendations of the NRCL. Hence, differences in positivity of the various test kits might not only be inherent to the kit itself but also the performance of the test.

It is evident that diagnostic test practices, including patient selection for testing, choice of test method and the performance of the diagnostic test influence test outcomes. From our dataset, heterogeneity between preferred diagnostic test methods can be observed. There is a high degree of uncertainty linked to physicians’ testing behaviours and also test performance in the laboratories, or rather the physicians’ adherence to existing guidelines. An assessment of current practices and the harmonisation across Switzerland could improve public health surveillance and decrease heterogeneity (e.g. of levels of underestimation) between regions.

### Limitations

For feasibility reasons, considering over 106 laboratories are either authorised or accredited in Switzerland, the analysis had to be limited to a selection of laboratories [37]. We chose to base the selection on the volume of notifications during the study period, therefore, favouring laboratories with the highest notification rates. With the 14 selected laboratories 54% of all notifications between 2007-2016 could be covered. We thus, believe that this may result in a negligible bias in our data.

## CONCLUSION

We found a stable positivity for *Legionella* spp. testing between 2007 and 2016 analysing the testing data of 14 Swiss diagnostic laboratories. There is a proportional increase in the number of cases identified in relation to the number of diagnostic tests performed. However, it is not clear why the number of tests performed more than doubled in the 10 year study period. The interpretation of the positivity curve and the implications on disease incidence can be vastly different depending on the reason for the increase in testing volume.

The assessment is further complicated, as large variation in positivity and test volume across the seven greater regions of Switzerland exists. We assume that these differences are only partly explained by differences in actual disease incidence; they seem to also stem from different CAP case management and diagnosis plans and represent different degrees of underestimation. The scarcity of data impedes evaluation of the different hypotheses. The diagnostic method applied greatly influences the test outcome. Culture-based methods, PCR and UATs perform differently and have their own limitations; particularly as in the case of the latter heating practices are suspected to vary.

The lack of national (or adherence to existing) guidelines, the heterogeneity of the diagnostic tests and in testing procedures applied, hampers the diagnosis of LD as well as comparison of data in a public health context. Therefore, diagnostic procedures should be harmonised across Switzerland to follow recommendations from the national reference centre for *Legionella*.

## Data Availability

The data that support the findings of this study are available from the corresponding author, DM, with the permission of the FOPH and the FSVO, upon reasonable request.

## COMPLIANCE WITH ETHICAL REQUIREMENTS

### Author contributions

CS and DM conceived and designed the study. Data collection and processing was performed by FF, with CS. FF conducted the analysis. FF, CS, VG and DM interpreted the results. FF wrote the first draft of the manuscript. All authors contributed to the revisions of the manuscript and approved the final version.

### Funding

This study was funded by the Swiss Federal Office of Public Health (FOPH, grant number 16.015253) and the Swiss Federal Food Safety and Veterinary Office (FSVO, grant number 307.2/2014/00158).

### Ethical approval

The study was conducted under the Epidemics Act (SR 818.101). The data, provided by laboratories, were anonymised for the analysis. Other data (e.g. notification data, population statistics) is publicly available from the FOPH or the Swiss Federal Statistical Office.

## ACKNOWLEDGMENTS

The authors thank Apolline Saucy (Swiss Tropical and Public Health Institute, Swiss TPH) for support during the data collection and data cleaning. Christian Schindler (Swiss TPH) and Jan Hattendorf (Swiss TPH) provided statistical advice. Various staff of the Federal Office of Public Health (FOPH) provided detailed insights to the Swiss surveillance system and information on the notification data. We specifically thank Mirjam Mäusezahl (FOPH) and Nicole Gysin (FOPH) for reviewing and commenting to the manuscript The authors much appreciate the support of the following laboratories providing data for the study: ADMed Microbiologie / Lienhard Reto (La Chaux-de-Fonds), EOLAB - Dipartimento di medicina di laboratorio, Ente Ospedaliero Cantonale (Bellinzona), Institut für Infektionskrankheiten (IFIK, Bern), Institut für Labormedizin soH AG (Solothurn), Institut für medizinische Mikrobiologie (IMM, Luzern), Kantonsspital Aarau AG (Aarau), Laboratoire de Bactériologie des HUG / Jacques Schrenzel (Geneva), Spitalzentrum Biel AG (Biel), Universitätsspital Basel / Adrian Egli (Basel), Viollier AG (Allschwil), Zentrum für Labormedizin (St. Gallen) and three other Swiss diagnostic laboratories. The Federal Office of Public Health and the Federal Food Safety and Veterinary Office are gratefully acknowledged for providing the funding and the framework for this study.

**ESM 1 Online Resource on repeated tests** Description of the definition of a disease episode of a *Legionella* spp. infection and the exclusion of repeated tests from the raw dataset by 14 Swiss laboratories (2007-2016)

- **Supplementary Fig 3 Different example scenarios based on the definition of disease episode to** exclude repeated tests for *Legionella* spp. in Switzerland, 2007-2016.

**ESM 2 Online Resource on serological tests** Descriptive analysis of the serological tests performed for Legionella spp. in Switzerland (2007-2016) as provided in the raw dataset by 14 Swiss laboratories and argumentation for their exclusion for further analysis

**ESM 3 Online Resource for additional figures**

- **Supplementary Fig 1 Seasonality in test volume and cases** The average and interquartile range (IQR) per calendar month of the total number of *Legionella* spp. tests and number of positive tests, Switzerland, 2007-2016
- **Supplementary Fig 2 Age distribution in test volume and positivity** a: Positivity of *Legionella* spp. testing by sex and age groups, Switzerland, 2007-2016. b: Number of *Legionella* spp. tests performed by sex and age groups in Switzerland (2007-2016) and permanent resident population in Switzerland (2016) by sex and age groups
- **Supplementary Fig 3 Correlation of laboratories and region** Correlation between the variables “greater region” and “laboratory” included in the *Legionella* spp. positivity study, Switzerland, 2007-2016

## Footnotes

1 Fischer FB, Saucy A, Schmutz C, Mäusezahl D. Do changes in EHEC diagnostics mislead interpretation of disease surveillance data in Switzerland? Time trends in positivity from 2007 to 2016. 2019. Accepted (*Eurosurveillance*).

